# Insights from an autism imaging biomarker challenge: promises and threats to biomarker discovery

**DOI:** 10.1101/2021.11.24.21266768

**Authors:** Nicolas Traut, Katja Heuer, Guillaume Lemaître, Anita Beggiato, David Germanaud, Monique Elmaleh, Alban Bethegnies, Laurent Bonnasse-Gahot, Weidong Cai, Stanislas Chambon, Freddy Cliquet, Ayoub Ghriss, Nicolas Guigui, Amicie de Pierrefeu, Meng Wang, Valentina Zantedeschi, Alexandre Boucaud, Joris van den Bossche, Balázs Kegl, Richard Delorme, Thomas Bourgeron, Roberto Toro, Gaël Varoquaux

## Abstract

MRI has been extensively used to identify anatomical and functional differences in Autism Spectrum Disorder (ASD). Yet, many of these findings have proven difficult to replicate because studies rely on small cohorts and are built on many complex, undisclosed, analytic choices. We conducted an international challenge to predict ASD diagnosis from MRI data, where we provided preprocessed anatomical and functional MRI data from > 2,000 individuals. Evaluation of the predictions was rigorously blinded. 146 challengers submitted prediction algorithms, which were evaluated at the end of the challenge using unseen data and an additional acquisition site. On the best algorithms, we studied the importance of MRI modalities, brain regions, and sample size. We found evidence that MRI could predict ASD diagnosis: the 10 best algorithms reliably predicted diagnosis with AUC∼0.80 – far superior to what can be currently obtained using genotyping data in cohorts 20-times larger. We observed that functional MRI was more important for prediction than anatomical MRI, and that increasing sample size steadily increased prediction accuracy, providing an efficient strategy to improve biomarkers. We also observed that despite a strong incentive to generalise to unseen data, model development on a given dataset faces the risk of overfitting: performing well in cross-validation on the data at hand, but not generalising. Finally, we were able to predict ASD diagnosis on an external sample added after the end of the challenge (EU-AIMS), although with a lower prediction accuracy (AUC=0.72). This indicates that despite being based on a large multisite cohort, our challenge still produced biomarkers fragile in the face of dataset shifts.

## Introduction

Autism Spectrum Disorder (ASD) is a life-long neurodevelopmental disorder which affects more than 1% of the population. Its severity differs vastly among individuals, however, they all share persistent deficits in social communication and restricted, repetitive and stereotyped behaviours. ASD is heritable, and influenced by common genetic variation as well as rare mutations (Krumm et al. 2015; Bourgeron 2015; Sandin et al. 2017; Weiner et al. 2017). Early intervention has a significant positive impact on the patient’s outcome, which makes early diagnosis a research priority (Dawson et al. 2010).

Magnetic resonance imaging (MRI) is an important tool to explore the brain of individuals with ASD: it is a widely available, fast, and non-invasive method to measure brain anatomy and function. By providing detailed measurements of an individual’s brain, MRI brings the promises of precision psychiatry, adapting therapy to patients (Insel 2014). But can MRI be used to characterise ASD in general? For more than 30 years, MRI studies have described anatomical and functional differences between individuals with ASD and unaffected controls: enlarged brain volume and cortical surface area (Piven et al. 1995; E. Courchesne et al. 2001), decreases in brain volume and neocortical thinning during adolescence and adulthood (Lange et al. 2015; Zielinski et al. 2014), smaller corpus callosum (Egaas et al. 1995; Wolff et al. 2015), abnormal cerebellar volume (Courchesne 1987; Hodge et al. 2010; Fatemi et al. 2012), and global and regional increases and decreases in functional connectivity (Just 2004; Belmonte 2004; Di Martino et al. 2014a; Cheng et al. 2017).

Many of these findings are, however, controversial and have proven difficult to replicate (Haar et al. 2016; Lefebvre et al. 2015; Traut et al. 2018; Picci et al. 2016; Mohammad-Rezazadeh et al. 2016). Most studies have relied on sample sizes far too small to reach reliable conclusions – sometimes just a few dozen subjects, and up to a few hundreds at most. They lack replication and reanalysis on independent data. This is particularly problematic because of the multitude of parameters involved in each analysis which could substantially alter the results (Carp 2012; Power et al. 2012; Poldrack et al. 2017): acquisition sequence, subject motion, software packages, pre-processing workflow, etc.

Rather than focusing on the detection of specific regional differences between cases and controls, brain-imaging features can be combined into a biomarker of ASD answering the question: can diagnostic status be inferred from MRI data? Machine-learning provides important techniques to build and characterise such biomarkers. Yet, machine-learning studies of ASD are most often based on the analysis of a single sample, without validation of the findings in an independent sample. The community recognises today that establishing the validity of a biomarker needs a fully independent assessment on new data, otherwise its accuracy cannot be trusted (Woo et al. 2017; Poldrack et al. 2019) as it may arise from overfitting, circular analysis (Kriegeskorte et al. 2009) or researchers’ degrees of freedom (Ioannidis 2005). This is particularly critical for machine learning approaches, where classifiers trained on data from one sample may be unable to generalise to additional samples (Ecker et al. 2015). Publication incentives lead researchers to seek and report the best prediction accuracy. For brain-imaging biomarkers of ASD, publications have reported accuracies above 95% (Bi et al. 2018). If that were true, the accuracy of those algorithms would be equivalent to the inter-rater reliability of clinical assessment by human experts (kappa=95%) which defines the gold-standard for discrimination of ASD versus other development disorders (Klin et al. 2000). But how trustworthy is the evaluation of biomarkers such as those in (Bi et al. 2018), given that the whole study – biomarker extraction and validation – was done on only 50 ASD patients and 42 non-ASD controls? Peer-review is not sufficient to assess the analytic choices as even minor variants lead to large differences in observed prediction accuracy, though these are unlikely to reveal true improvements (Varoquaux 2018).

To ground solid conclusions on ASD neuroimaging, several international consortia have been constituted such as ABIDE (Autism Brain Imaging Data Exchange, Di Martino et al. 2014b; Di Martino et al. 2017), EU-AIMS (European Autism Interventions - A Multicentre Study for Developing New Medications, Murphy and Spooren 2012) or the IBIS Network (Infant Brain Imaging Study, Hazlett et al. 2012), increasing sample sizes through data sharing. They extend the amount and quality of the data collected through harmonisation efforts. Recent analysis across cohorts has shown that ASD is significantly associated with changes in functional connectivity (Holiga et al. 2019). But are these changes large enough to ground reliable prediction to new sites, despite heterogeneity in imaging techniques and populations recruited?

The study we present built upon these large cohorts, and framed the extraction of biomarkers as an open, international challenge to predict ASD from the largest MRI dataset currently available – more than 2,000 individuals. A data-science prediction challenge of this type can provide conclusive evidence on the ability of MRI to detect ASD, because it is based on a blind evaluation of the results in addition to relying on a large sample. Furthermore, it isolates the development of the analysis pipeline from its evaluation. Challengers did not have access to validation data, which allowed us to test the ability of the algorithms to generalise to unseen data, including data acquired in different centres.

## Materials and methods

### Brain imaging dataset

The brain imaging dataset combined data from the public Autism Brain Imaging Data Exchange (ABIDE) I and II datasets (Di Martino et al. 2014a; Di Martino et al. 2017) and an unpublished dataset from the Robert Debré Hospital (RDB) in Paris, France (see Supplemental Table 1 for additional demographic information for the RDB site). ABIDE provides open access to functional and anatomical MRI data for 2,156 subjects. The RDB dataset contained data from 247 subjects, 56 of whom were also part of the ABIDE II project: we excluded these duplicate subjects from the private dataset. With the exception of data coming from the RDB centre which was acquired in a 1.5 Tesla scanner, all MRI data was acquired in 3 Tesla scanners. In all sites subjects were diagnosed using standard ADI/ADOS tools to support clinical assessment. Most subjects had a full IQ>75.

We extracted anatomical features from the anatomical MRI: regional brain volumes, cortical thickness, and surface area; and extracted time-series signals from the resting-state functional MRI. To derive these individual measurements, the data were processed with standard neuroimaging tools: Freesurfer (Fischl et al. 1999), FSL (Woolrich et al. 2009), and AFNI (Cox 1996). We split the total dataset into public and private datasets, aiming at balancing the age and sex distributions (Table 1 and Fig. 1 provide demographic information for the public and private datasets). A total of 2,117 subjects were included: 947 ASD and 1,170 controls. We did not exclude subjects based on quality control, but provided challengers with quality control scores obtained from visual inspection by 3 experts. The public dataset contained data from 1,150 subjects: 549 ASD and 601 controls. The private dataset contained data from 967 subjects: 398 ASD and 569 controls. The public dataset was a subset of ABIDE I and II, while the private dataset combined data from ABIDE I, II and RDB. The dataset should capture well the clinical and methodological heterogeneity of ASD neuroimaging. The ABIDE dataset was collected by 24 different centres worldwide, and spans an age range from 5 to 64 years old (median 13.8 years old). All subjects had intellectual quotients within the normal range (97% have full IQ>80), 80% of patients and 90% of controls were right-handed. The cohort was composed of 80% males.

**Table 1.** Demographic information. M: male, F: female. Values for age and full scale IQ are summarised in the form mean ± standard deviation (min - max). Full scale IQ was not available for every individual.

| Dataset | Variable | Control | ASD |
| --- | --- | --- | --- |
| Public set (ABIDE) | Sex | 449 M, 152 F | 471 M, 78 F |
| | Age (years) | 16.89 ± 9.46 (5.89 - 56.2) | 17.17 ± 10.5 (5.13 - 64.0) |
| | Full scale IQ | 112.92 ± 13.06 (71.0 - 149.0) | 105.85 ± 16.66 (61.0 - 149.0) |
| Private set (ABIDE+RDB) | Sex | 387 M, 182 F | 351 M, 47 F |
| | Age (years) | 20.61 ± 13.45 (4.0 - 70.8) | 14.56 ± 6.62 (4.7 - 45.0) |
| | Full scale IQ | 112.39 ± 12.45 (40.0 - 142.0) | 104.62 ± 18.22 (41.0 - 149.0) |
| Replication set (EU-AIMS) | Sex | 174 M, 94 F | 263 M, 97 F |
| | Age (years) | 16.95 ± 5.7 (6.89 - 30.98) | 16.63 ± 5.48 (7.08 - 30.33) |
| | Full scale IQ | 103.55 ± 19.28 (50.0 - 142.0) | 98.64 ± 19.4 (54.97 - 148.0) |

**Figure 1.**
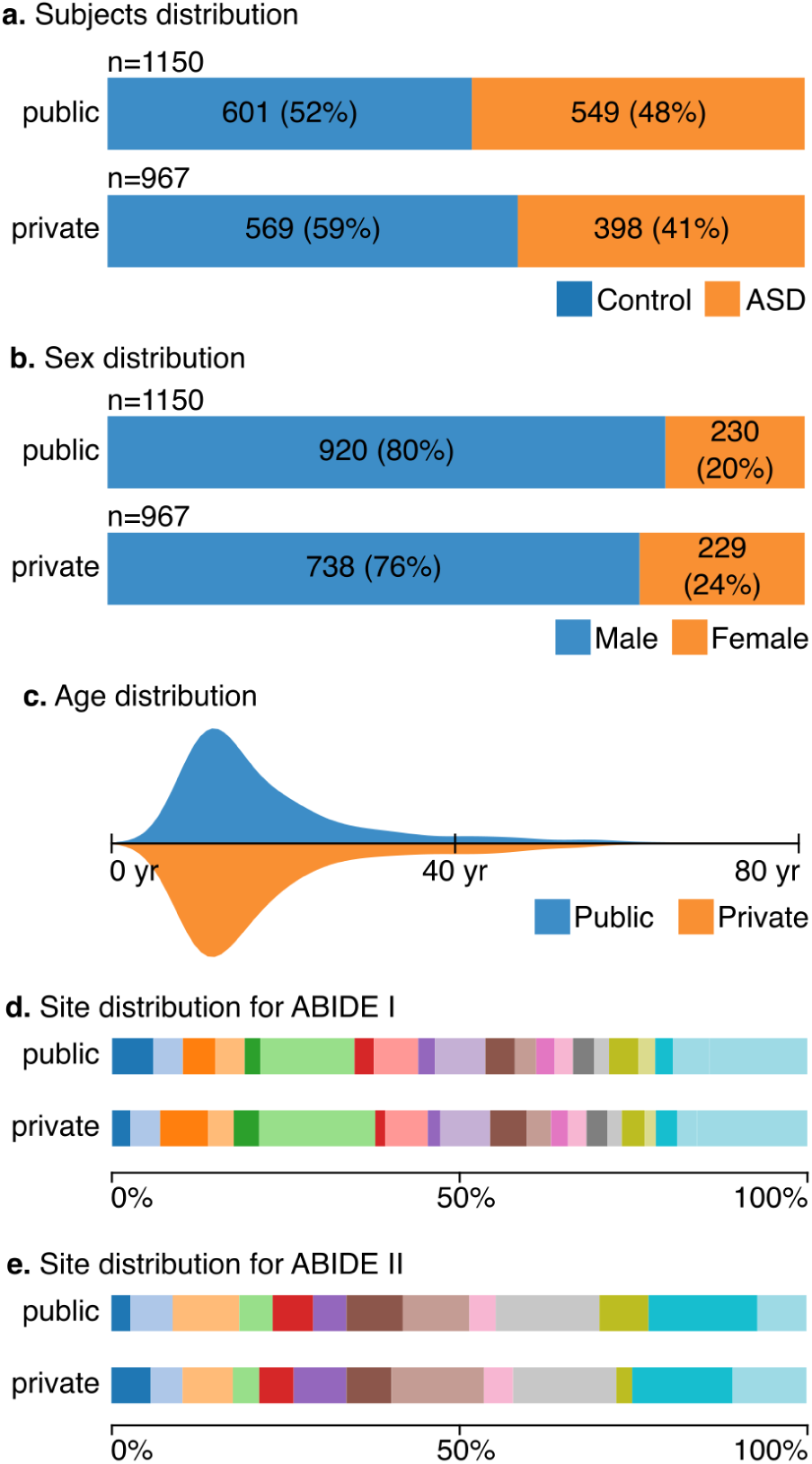
Subjects, sex, age, and site distributions. The distributions of the number of cases and controls, their sex ratio, age and scanning site were similar in the public and private datasets.

This diversity should allow classification algorithms to generalise, preventing them from specialising in a particular type of data or age range. Further demographic information for the ABIDE I and II datasets is provided in their corresponding publications (Di Martino et al. 2014a; Di Martino et al. 2017).

### Study design

#### a data-science prediction challenge

##### Description of the challenge

We launched the challenge inviting data scientists to submit algorithms to predict ASD diagnostic from provided MRI data. The challenge lasted 3 months and attracted 146 challengers. We awarded money prizes to the 10 best challengers. These were determined after the closure of the competition, by assessing how well the algorithms would predict ASD diagnostic in a private, unseen dataset.

The ABIDE data can be openly distributed, which enabled us to provide a rich dataset on which challengers could tune their algorithms. To facilitate access to the data, we provided challengers with a “starting kit” giving a proof-of-concept predictive model on the data (extracted on standard brain atlases). Challengers were then able to develop their own prediction algorithms, which they submitted using a Web interface. The code was executed on our central server. The challengers never had access to the private dataset. They used machine-learning techniques tuned on the public dataset and submitted the corresponding code to the central server which evaluated them in the private dataset.

To measure the quality of the predictions, we used a standard metric: the area under the receiver operating characteristic curve (ROC-AUC). This measure summarises various detection tradeoffs, for example, favouring few false negatives to the cost of false positives in the case of a screening study, or the converse, in the case of a confirmatory study. Prediction at chance level gives an AUC of 0.5, while perfect prediction gives an AUC of 1.

##### Statististical analysis strategy

After the closure of the challenge, we analysed the 10 best submissions to understand the factors driving their predictions. Considering that these machine-learning algorithms captured the best possible predictive biomarkers given our brain-imaging cohort, we varied the data on which they were applied. Each time, we fit the algorithms anew on the data, to extract the corresponding biomarkers, and measured the prediction accuracy on the private dataset. First, we applied them to different imaging modalities: only functional MRI, or only anatomical MRI. Second, we varied the number of available subjects, to measure the importance of the sample size. Finally, we investigated the importance of different brain regions by removing those that appeared as most discriminative and attempting to extract biomarkers from the rest of the data. To compute which regions were the most discriminative, we used the absolute value of the model’s coefficients when the algorithm used a linear model, and the feature importance when the algorithm used a random forest. We obtained a region-level summary of functional-connectivity biomarkers by associating to every region the sum of the importance of its connections, a measure of node strength. Region-level importance was then turned into a brain map characterising the spatial distribution of the discriminant information.

#### MRI preprocessing and signal extraction

##### Anatomical MRI

Anatomical MRI was preprocessed and segmented using FreeSurfer v6.0. We extracted three kinds of anatomical features: (i) mean regional cortical thickness, (ii) cortical surface area of regions parcellated with the Desikan-Killiany Atlas (Desikan et al. 2006), and (iii) volumes of subcortical regions segmented with the FreeSurfer atlas (Fischl et al. 2002).

##### Resting-state functional MRI

Resting-state functional MRI captures brain activity and functional connectivity. It is typically studied via a functional-connectome: a matrix capturing interactions between brain regions. We provided time series extracted on a variety of atlases after standard fMRI preprocessing using the pipeline from the FC1000 project (which includes slice-time interpolation, motion correction, coregistration to anatomic data, normalisation to template space). The brain parcellations and atlases used were: (i) BASC parcellations with 64, 122, and 197 regions (Bellec et al. 2010); (ii) Ncuts parcellations (Craddock et al. 2012); (iii) Harvard-Oxford anatomical parcellations; (iv) MSDL functional atlas (Varoquaux et al. 2011); and (v) Power atlas (Power et al. 2011).

#### Challenge organisation

##### Organisation of the challenge

We launched the challenge on May 1 ^st^ 2018 and closed it on July 1 ^st^ 2018. The challenge attracted about 146 participants accounting for a total of about 720 submissions. We awarded money prizes to the 10 best challengers. Challengers were ranked based on the ROC-AUC score of their submission computed on the private dataset. We framed the challenge problem by providing: (i) a public dataset, (ii) a standard way to assess the submission, and (iii) a starting kit. For this purpose, we used the RAMP (Rapid Analytics and Model Prototyping) workbench. Participants submitted their solutions on the RAMP website (https://ramp.studio). During the challenge we provided to the participants the cross-validated score computed on the public dataset. At the end of the challenge, we asked each participant to select a single submission. This submission was trained (fitted) on the public dataset and evaluated on the private dataset hidden from the participants. Participants were ranked based on the score of these submissions computed on the private dataset.

##### Challenge platform

RAMP is an online data science tool used to organise challenges. RAMP enables us to easily compare and reproduce predictive experiments. It can be used on the user’s computer or online: the former is for developing predictive models while the latter assesses their predictive accuracy. A RAMP “starting kit” is a placeholder where we define the data-science problem: we provide the datasets, the metric, and the model validation technique. Participants can focus on the development of their machine-learning predictive model. We also provide examples to help participants understand the challenge. The RAMP website was used to evaluate the solutions of the participants (i.e., predictive models): participants submit their code and the website trains (fits) and evaluates them. We deployed, trained, and tested the full workflow on Amazon Web Services. Note that participants can also train and test their models locally. However, they only have access to the public dataset to test their models. We rely on Python and its rich ecosystem. Participants were free to use any Python library to build their machine learning model.

### Data and code availability

The public data, the code of the ten best submissions, the MRI preprocessing scripts and the scripts used to generate the figures are available at https://github.com/neuroanatomy/autism-challenge.

## Results

### Participation to the challenge

We received 589 submissions from 61 teams. To assess external validity of the biomarkers (Steyerberg and Harrell 2016), the private dataset contained >200 subjects not included in ABIDE from the Robert Debré Hospital in Paris, France (RDB). We selected the 10 submissions which performed the best (taking only one submission per team) as the winning ones, and gave money prizes to the submitters. After close inspection we discovered that we forgot to remove 56 subjects of the RDB dataset which were also in the ABIDE dataset. For the post-hoc analyses presented here, we removed those duplicated subjects from the private dataset to avoid artificially inflating the prediction score. The ranking between the 10 best submissions remained relatively stable after the removal of the duplicate subjects (Spearman correlation: rs = 0.7697, p (2-tailed) = 0.00922) (see Supplemental Table 2 for a detail of the performance of each submission before and after the removal of duplicated subjects).

### The combination of the 10 best models provided a good predictor of ASD diagnosis

We combined the 10 best models using a blending approach (Caruana et al. 2004) to produce a probability for ASD diagnosis for each individual. These probabilities were in general higher for patients than for controls (Fig. 2a). The receiver operating curve (ROC, Fig. 2b, Supplemental Methods M1) represents the quality of these predictions for different tradeoffs of sensitivity and specificity, which can be summarised by the area under the curve (AUC). The combined predictor reached an AUC of 0.80, which is considered as a good discrimination level (Hosmer and Lemeshow 2000). Used as a screening test, the predictor would correctly detect 88% of the individuals with ASD at the cost of misclassifying 50% of controls as patients. Used as a confirmatory diagnostic test, the predictor would detect 25% of the individuals with ASD and only 3% of controls would be misclassified as patients. Predictions were also good (even slightly better) on the RDB subjects which were absent from the ABIDE dataset (median AUC=0.809 versus AUC=0.768 – Fig. 2d).

**Figure 2.**
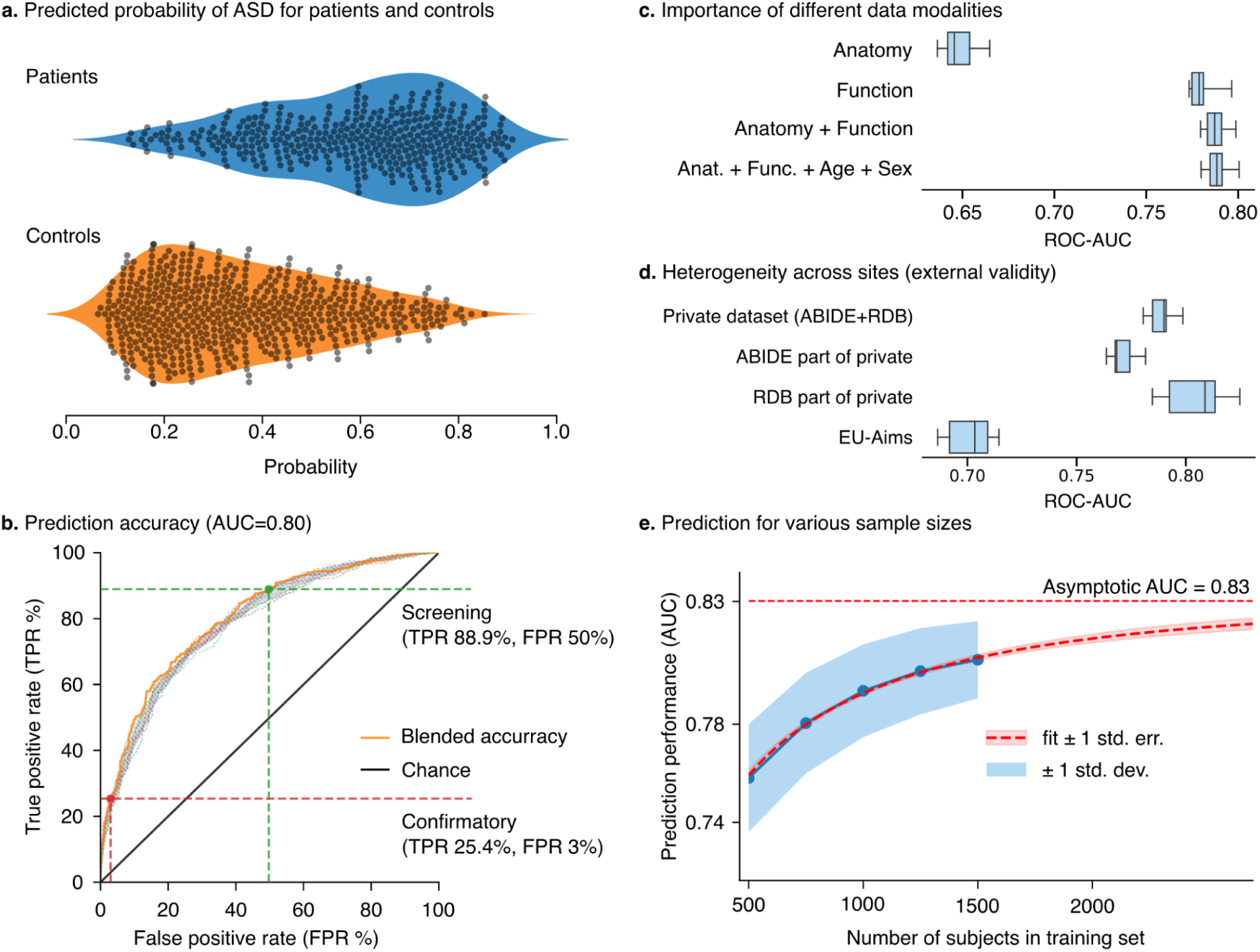
Performance analysis using the 10 best submissions of the challenge. (a) Predicted probability for patients and controls to be classified with ASD. (b) Prediction accuracy obtained by combining all available data modalities (anatomical and functional MRI, age, and sex). Using the biomarkers for screening purposes led to a True Positive Rate (TPR) of 88.9% for a False Positive Rate (FPR) of 50%. Enforcing a low FPR (3%) to make a confirmatory analysis led to a True Positive Rate of 25.4%. (c) Importance of the data modalities to predict ASD. Functional MRI provided a higher discriminative power than the other available data (population indicators and anatomical MRI data). (d) Data heterogeneity across sites was not a roadblock: methods generalised to data from new sites, unseen during training. (e) Prediction accuracy by varying the number of subjects in training. Prediction accuracy did not reach a plateau with the current number of subjects available. Increasing the number of subjects improves the discriminative power of the methods. The current trend suggests an increase of prediction performance to 0.83 for a dataset of 10,000 subjects.

After the challenge, we used an additional dataset from the EU-AIMS project to evaluate the ability of the blended predictor to generalise. Performance on this additional data was slightly worse than on our external site (AUC=0.721), revealing that the predictor was still fragile to distribution shifts despite having been extracted from multiple sites. Demographic information for this dataset can be found in Table 1; the most pronounced difference between EU-AIMS and our study cohort is on the IQ of participants and we speculate that this difference might drive the prediction performance loss.

### Functional MRI had the strongest impact on prediction accuracy

Studying separately the prediction accuracy obtained for each imaging modality showed that functional MRI contributed more to prediction than anatomical MRI (AUC=0.79 using only functional MRI, versus AUC=0.66 using only anatomical MRI, Fig. 2c). Incorporating age and sex information only had a small influence on prediction accuracy (increasing AUC to 0.80). See Supplemental Methods M2 for additional information.

### Further increases in sample size should lead to increased prediction accuracy

The analysis of the learning curve showed that prediction accuracy was not reaching a plateau and should keep improving by increasing the number of subjects (Fig. 2e). Extrapolating on this increase, we estimate that prediction accuracy could reach an AUC=0.83 if a dataset of 10,000 subjects were available (see Supplemental Methods M3 for additional information).

### Regions distributed over the entire brain contributed to prediction

We analysed the functional MRI biomarkers to highlight the most discriminative brain regions. For this, we ranked brain regions by their importance in the 10 best models. Overall, prediction relied on regions distributed over the entire brain, with regions around the precuneus appearing to be the most important (Fig. 3a). We assessed the relative importance of the different brain regions by progressively removing those that contributed the most and extracting new biomarkers. Prediction accuracy remained high even after removing up to 50% of the most important brain regions (see Fig. 3b). This suggests that the biomarkers captured information distributed over the entire brain (see Supplemental Methods M4 for additional information).

**Figure 3.**
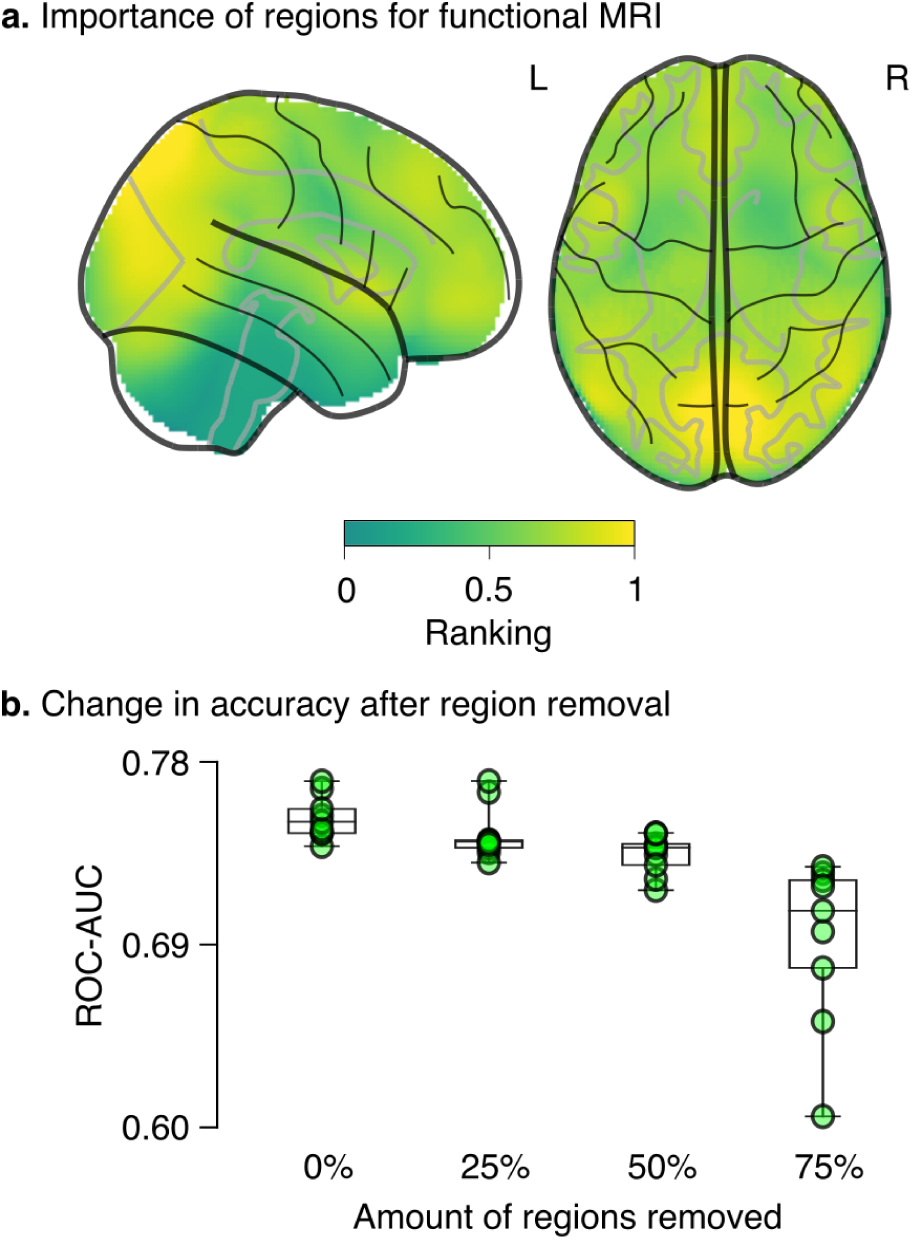
Regions important for the functional-MRI biomarker. (a) statistical map giving the ranking of regions’ contribution to predictions from the functional data; (b) decrease in performance by removing the most important regions.

## Discussion and conclusions

### MRI can provide reliable biomarkers for ASD

The results of our challenge conclusively demonstrate that MRI is a powerful and reliable method to study ASD. Interestingly, it is the size of the cohort – not interindividual heterogeneity – that is the main factor limiting prediction accuracy. This can be seen from the fact that aggregating data across sites with different recruitment policies led to a steady increase in prediction accuracy. Increasing sample size is so far the best strategy for achieving further progress: better predictive power and better spatial localisation of the biomarkers. We project that a population of 10,000 individuals should allow us to reach the maximum prediction accuracy that a simple case-control design can achieve. At this stage, the adoption of a dimensional approach to model ASD should lead to further improvements in the prediction of clinical status.

The results of our challenge suggest that MRI provides an important source of information for the study of ASD, complementary to that obtained, for example, through genetic exploration. Recent reports show that polygenic risk scores (PRS), aiming to predict case-control status from genome-wide common variants (Purcell et al. 2009), can explain 2.45% of the risk variance on the observed scale (Nagelkerke pseudo-R ^2^) in a group of 13,076 ASD cases and 22,664 controls (Grove et al. 2019). Considering a disease prevalence of 1.2%, PRS should capture 1.13% of the risk variance of ASD on the liability scale (Lee et al. 2012). As a matter of comparison, the AUC of 0.80 that we obtain from MRI corresponds to an R ^2^ of 19.1% on a liability scale with the same disease prevalence (Wray et al. 2010). We can expect that larger genetic samples for ASD should increase prediction accuracy, as the proportion of risk variance captured by common genetic variants is estimated to be 11.8% on the liability scale (Grove et al. 2019). The important gap with prediction accuracy could be explained by differences in inclusion criteria, as large scale genetics studies include subjects with a wider range of intellectual ability. However, it could also be the case that the effects of common genetic variants are highly diluted across the genome, and therefore difficult to estimate individually. It may be informative to consider the case of schizophrenia – a neurodevelopmental disorder with overlapping genetic aetiology. In schizophrenia, PRS obtained from approximately 35,000 cases and 45,000 controls captured ∼15% of the risk variance on the observed scale (Nagelkerke pseudo-R ^2^) and ∼7% on the liability scale with an AUC of 0.70 (Schizophrenia Working Group of the Psychiatric Genomics Consortium 2014). Our MRI-based prediction of ASD provides a higher accuracy than what PRS can provide for schizophrenia, with a much smaller cohort. MRI-based prediction may have the additional benefit of providing longitudinal information which could be important to track disease progression. In combination, genetic and MRI information could provide a powerful tool to predict and understand ASD risk.

### Classic machine learning methods provided the best results

The starting kit we presented transformed the resting state fMRI time series from the MSDL atlas to a tangent correlation matrix (Varoquaux et al. 2010) and regressed stacked values from the matrix and anatomical values with a L1-penalised logistic regression. The starting kit combined the predictions from the functional and anatomical MRI with a meta-classifier based on a logistic regression. The transformation from the functional MRI time series to a tangent correlation matrix was adopted by all submissions; however, the choice of atlas was variable. Seven submissions used time series from several atlases, while three submissions used only one atlas. In addition to the transformation to a tangent correlation matrix, one submission transformed to a standard correlation matrix and another transformed to a partial correlation matrix. One submission performed dimensionality reduction using a principal component analysis (PCA). Six submissions used logistic regression as a first layer predictor, two used linear c-support vector classification (SVC) and the two others a combination of different methods. Three submissions obtained directly a single prediction whereas the remaining seven obtained several predictions, which were combined using a logistic regression for five of them, average for one them, and a majority vote for the last one.

Challengers used a variety of methods, which included machine-learning techniques ranging from simple logistic regressions to complex graph-convolutional deep-learning models. Inspection of the individual scores showed that deep-learning techniques displayed strong overfits: good performance on the public dataset but poor generalisation to the private dataset. On the opposite, many algorithms which used simpler approaches had a stable prediction performance when applied to new data, displaying prediction accuracies between AUC=0.7 and 0.8 which did generalise to the private dataset. The submissions which led to reliable predictions had some methodology in common. In particular, they used linear support vector machines or logistic regression to compute the combination of the different signals – anatomical and functional – that best discriminated patients from controls. In addition, several submissions combined signals from multiple atlases using a stacking strategy (Wolpert 1992).

### Most improvement came from using atlases with larger numbers of regions

To speed up the computation, we presented a starting kit with the smaller atlas for resting state functional MRI. We saw that switching to larger atlases considerably increased the AUC: switching from the MSDL atlas (39 regions) to the craddock_scorr_mean atlas (249 regions) increased the AUC of the functional starting kit from 0.655 to 0.778 and the AUC of combined anatomy and functional from 0.716 to 0.790. The best submission we had obtained an AUC of 0.799 – an increase of less than 0.009 (see Supplement Table 2 for the performances of the different evolutions of the starting kit and the 10 best submissions).

Although we talk about functional and anatomical MRI in general, it is important to keep in mind that the variables we provided do not exhaust all possible functional and anatomical data. Though we used fairly standard preprocessing and feature-extraction choices, functional variables only include resting state time series, and anatomical variables only include surface, thickness and volume of different regions. The reason is that these are the types of measurements which can be acquired reliably in large populations using standard processing pipelines. Including additional functional and anatomical variables, such structural correlations or measurements obtained from diffusion-weighted imaging or T1/T2 ratios, may contribute to increasing the prediction power.

### Evaluation without a blind validation set is at risk of severe optimism

Our challenge highlights that a trustworthy development of biomarkers must include the evaluation on a new dataset, blinded to the analysts. In our challenge, participants had no interest in developing algorithms that would give optimistic results on the public dataset, as they knew that they would be evaluated on the private dataset. Nevertheless, comparing the prediction quality on the public dataset to that on the private dataset (Figs. 4 and 5) clearly showed that the best predictions on the public dataset were too good to be true, and did not carry over to the unseen, private dataset. This is likely because the challengers made their analytic choices or trained hyperparameters to improve the prediction score that they measured on the public dataset (Fig. 4). As a result, they obtained seemingly-excellent predictions, but which likely relied on noise in the public data and did not generalise to the private dataset. Indeed, the techniques used in machine-learning to measure prediction performance, such as cross-validation, are not completely robust to systematic exploration of analytic choices (Varoquaux 2018): To fully trust the prediction accuracy of biomarkers, these must be validated externally, as in our challenge. However, when testing external validity on a third, completely new data (EU-AIMS, see Fig. 5), the ranking of the models stayed roughly similar as on the private dataset, a pattern already reported on kaggle challenges (Roelofs et al. 2019). The blind assessment was necessary to select methods that extract good ASD biomarkers, and to reliably quantify prediction accuracy.

**Figure 4.**
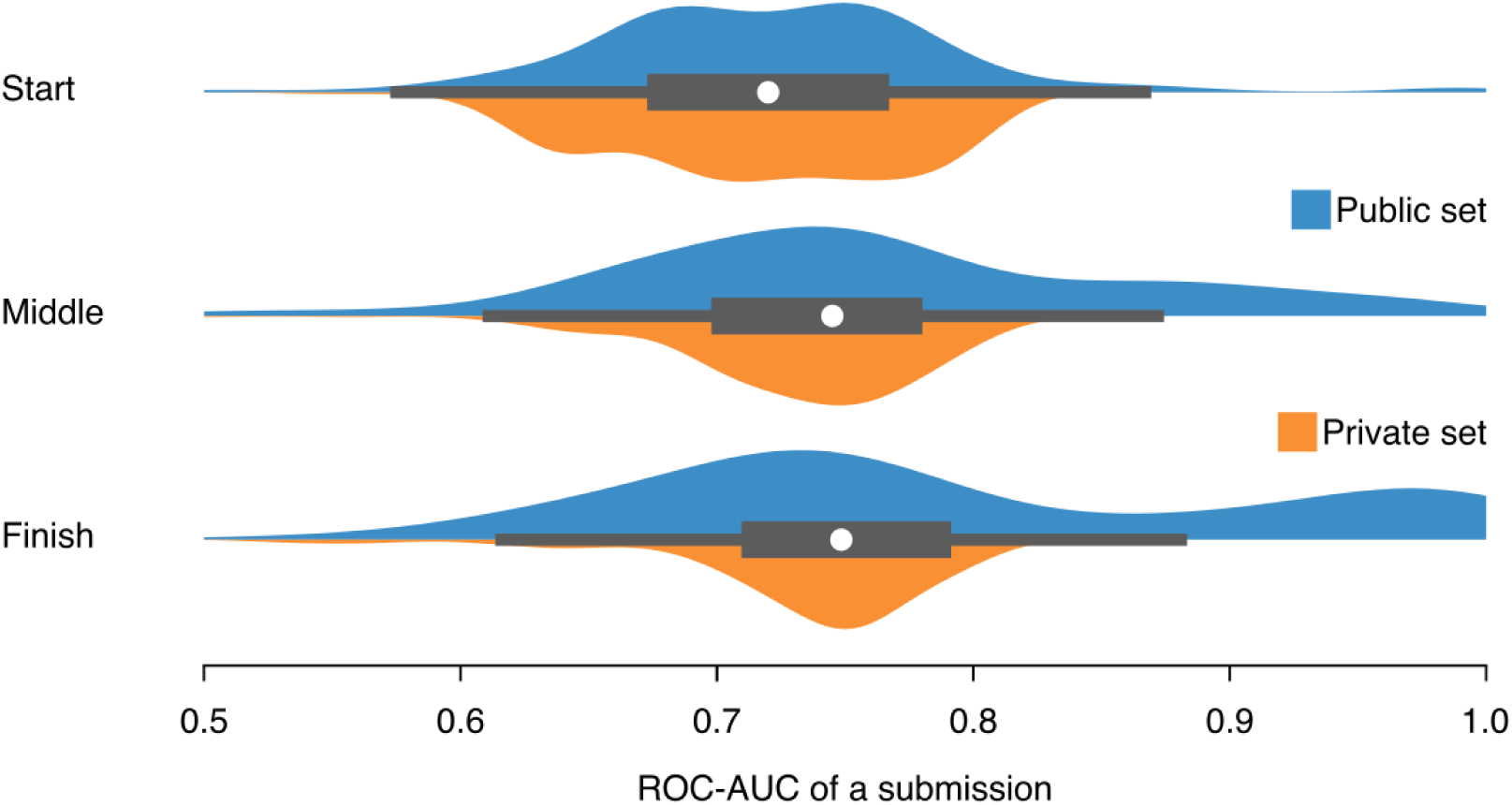
Score during the competition. The three different rows show prediction scores at the start of the challenge in May, at the middle of the challenge in June, and at the end of the challenge in July. The evolution of the scores of all submissions on the public and private datasets suggest that participants’ work led mostly to increased prediction performance in the public dataset without comparable increases in the private dataset.

**Figure 5.**
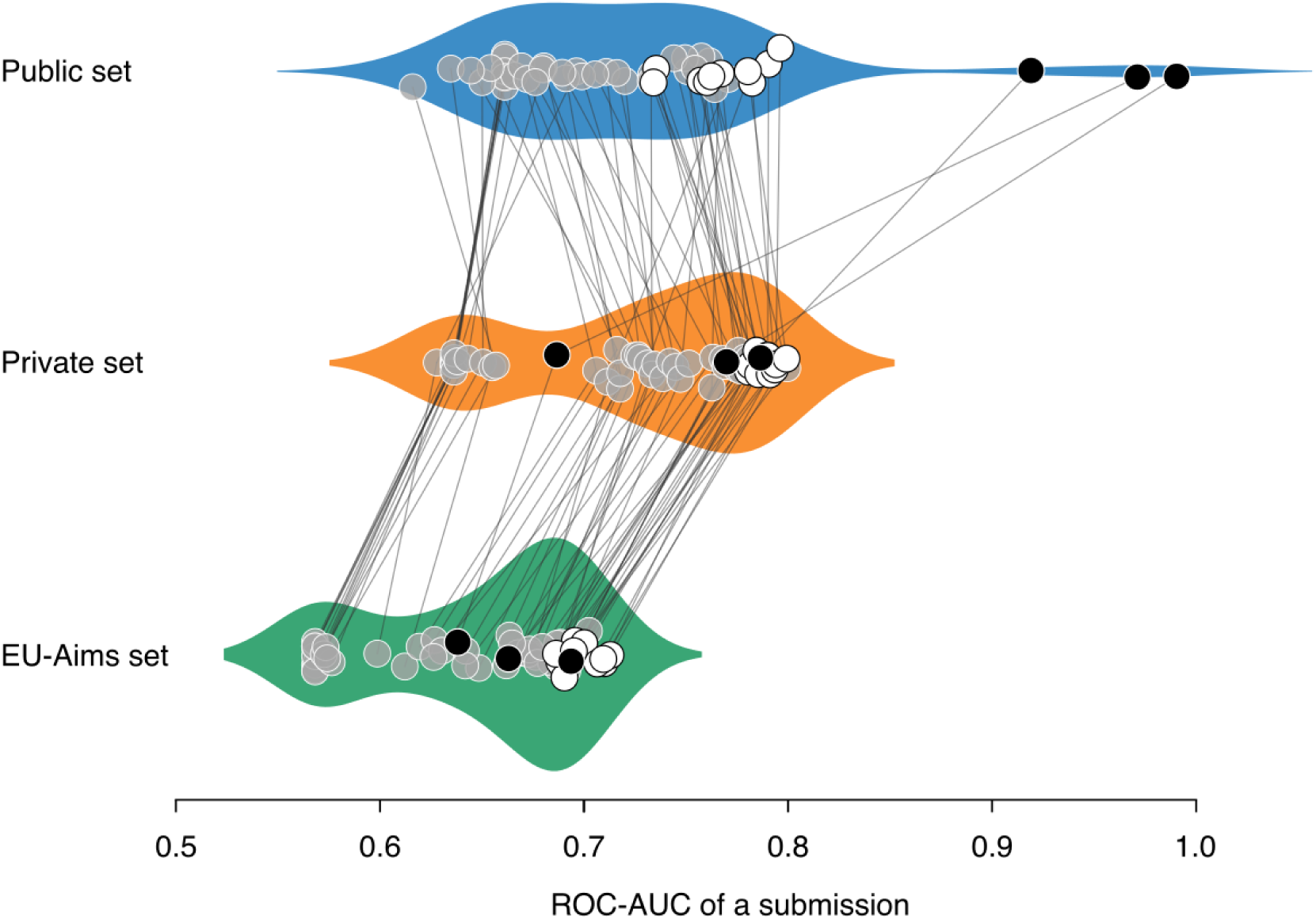
Excellent performance on the public dataset was misleading. The score of each final submission to the challenge is represented as a point on the public and the private dataset. The prediction on the public dataset was measured with 8 random splits (with 20% of the data left out), the standard machine-learning procedure to measure prediction performance. Extreme prediction scores in the public dataset (AUC>0.8) did not generalise to the private dataset. By contrast, algorithms with conservative scores (0.6<AUC<0.8) did predict the private dataset equally well.

### Better biomarkers bring new opportunities

Robust predictive MRI biomarkers open several opportunities for clinical and scientific research: Predictive MRI biomarkers enable longitudinal follow-ups and prospective epidemiology. Infants at risk of ASD could be scanned longitudinally, which could allow us to develop early biomarkers useful when behaviour is not a sufficient basis for diagnosis. Collecting and sharing brain-imaging data for 10,000 individuals at risk of ASD is well within the reach of a community effort. Such a large number would enable a substantial hold-out sample, and hence precise characterisation of the biomarkers. As predictive biomarkers reach an excellent prediction accuracy, the hope is that they will narrow down on the discriminant information. Hence, increases in prediction performance will also reveal more precise information on the neural correlates of ASD.

## Data Availability

Data for the public dataset are available online at https://github.com/neuroanatomy/autism-challenge

## Acknowledgements

We are grateful for the Paris-Saclay Center for Data Science for supporting this research.

