## Supplemental material for "Insights from an autism imaging biomarker challenge: promises and threats to biomarker discovery"

#### Supplemental tables

| *Age (in years)* | | | | |
|---|---|---|---|---|
| **Group** | **Number of subjects** | **Mean (Min, Max)** | **Median** | **Standard deviation** |
| All | 247 | 28.8 (4.0, 70.8) | 27.9 | 15.4 |
| ASD | 49 | 14.2 (4.7, 43.7) | 13.0 | 8.1 |
| Control | 198 | 32.4 (4.0, 70.8) | 34.4 | 14.7 |
| *Sex* | | | | |
| **Group** | **Female** | **Male** | | |
| All | 123 | 124 | | |
| ASD | 10 | 39 | | |
| Control | 113 | 85 | | |
| *Cognitive level* | | | | |
| **Group** | **Normal** | **Borderline** | **Delay** | |
| All | 227 | 10 | 9 | |
| ASD | 34 | 6 | 8 | |
| Control | 193 | 4 | 1 | |
| *Family status* | | | | |
| **Group** | **Proband** | **Relative** | | |
| All | 107 | 140 | | |
| ASD | 43 | 6 | | |
| Control | 64 | 134 | | |

**Supplemental Table 1**. Demographic information for subjects recruited at Robert Debré Hospital. Cognitive level was assessed by clinical judgement supported by a series of instruments: Raven's progressive matrices, WISC IV, WISC III, WPPSI IV, WAIS III, EDEI (*Échelles Différentielles d'Efficience Intellectuelle*).

| Submission | AUC orig | AUC bagged orig | AUC fixed | AUC bagged fixed | AUC EU-AIMS | AUC bagged EU-AIMS |
|---|---|---|---|---|---|---|
| **ayoub.ghriss_original** | 0.801197 | 0.805760 | 0.796429 | 0.798651 | 0.714034 | 0.711194 |
| **Slasnista_original** | 0.795106 | 0.801357 | 0.787399 | 0.793334 | 0.706893 | 0.706862 |
| **pearrr_original** | 0.794351 | 0.797825 | 0.788848 | 0.790119 | 0.707867 | 0.706551 |
| **lbg_original** | 0.791209 | 0.797763 | 0.788627 | 0.790402 | 0.703990 | 0.714438 |
| **amicie_original** | 0.779874 | 0.796618 | 0.765956 | 0.785443 | 0.682629 | 0.700218 |
| **mk_original** | 0.756596 | 0.796342 | 0.760887 | 0.791139 | 0.673974 | 0.686370 |
| **combine_anatomy_functional_craddock_scorr_mean** | 0.753364 | 0.795636 | 0.757231 | 0.790437 | 0.671828 | 0.686370 |
| **wwwwmmmm_original** | 0.783972 | 0.795468 | 0.781968 | 0.781133 | 0.707680 | 0.710106 |
| **nguigui_original** | 0.788383 | 0.794030 | 0.777799 | 0.780515 | 0.694714 | 0.695636 |
| **abethe_original** | 0.787582 | 0.793443 | 0.784785 | 0.789868 | 0.695543 | 0.690547 |
| **vzantedeschi_original** | 0.795020 | 0.793385 | 0.781464 | 0.784547 | 0.680079 | 0.688091 |
| **combine_anatomy_functional_basc197** | 0.752686 | 0.791159 | 0.755915 | 0.784114 | 0.675891 | 0.693688 |
| **starting_kit_functional_craddock_scorr_mean** | 0.781660 | 0.789418 | 0.771476 | 0.777848 | 0.679353 | 0.682577 |
| **starting_kit_functional_basc197** | 0.777426 | 0.783603 | 0.766327 | 0.771118 | 0.690153 | 0.692682 |
| **combine_anatomy_functional_basc122** | 0.747597 | 0.777787 | 0.745860 | 0.772271 | 0.679281 | 0.697450 |
| **combine_anatomy_functional_power_2011** | 0.736944 | 0.769302 | 0.737250 | 0.766870 | 0.668180 | 0.674420 |
| **starting_kit_functional_basc122** | 0.751380 | 0.761668 | 0.740279 | 0.749684 | 0.685634 | 0.693501 |
| **starting_kit_functional_power_2011** | 0.752612 | 0.759257 | 0.742173 | 0.746006 | 0.662303 | 0.665444 |
| **combine_anatomy_functional_basc064** | 0.709654 | 0.745466 | 0.725208 | 0.744050 | 0.648062 | 0.674274 |
| **combine_anatomy_functional_harvard_oxford_cort_prob_2mm** | 0.683723 | 0.742870 | 0.703743 | 0.738724 | 0.583022 | 0.626430 |
| **combine_anatomy_functional** | 0.653282 | 0.719962 | 0.689449 | 0.716266 | 0.583167 | 0.626835 |
| **starting_kit_functional_basc064** | 0.706216 | 0.717650 | 0.698051 | 0.711572 | 0.643968 | 0.650301 |
| **starting_kit_functional_harvard_oxford_cort_prob_2mm** | 0.676380 | 0.701900 | 0.668024 | 0.684998 | 0.572087 | 0.587883 |
| **starting_kit_functional** | 0.644921 | 0.671878 | 0.640364 | 0.655121 | 0.605338 | 0.612365 |
| **starting_kit_anatomy** | 0.635401 | 0.636341 | 0.630499 | 0.636385 | 0.573653 | 0.568377 |

**Supplemental Table 2**. Performance of ten best submissions and starting kit derivatives on the different datasets. **AUC orig**: AUC on the original private dataset, **AUC bagged orig**: bagged AUC on the original private dataset, **AUC fixed**: AUC on the fixed private dataset, **AUC bagged fixed**: bagged AUC on the fixed private dataset, **AUC EU-AIMS**: AUC on the EU-AIMS dataset, **AUC bagged EU-AIMS**: bagged AUC on the EU-AIMS dataset.

### Supplemental methods

*M1. ROC curve analysis*

A ROC curve (Fig. 2b) characterises the prediction accuracy of a binary classifier. Each point of the ROC curve shows the true positive rate against the false positive rate at a given discriminative threshold. For ASD detection, the true positive rate is the ratio of patients with ASD correctly identified as such. The false positive rate is the ratio of unaffected subjects wrongly identified as ASD patients. For each subject, a classifier outputs the probability of this subject to have ASD. Thus, the ROC curve is built by thresholding at different probabilities.

*M2. Effect of the input modality*

We studied the influence of each data modality on the prediction. For this, we changed the input data for each of the 10 best models. We trained and tested each algorithm using only anatomical features, only functional features, and the combination of the two. Additionally, we included another trial where we added age and sex information to the brain-imaging data. Figure 2c shows the prediction accuracy for each of these experiments. Algorithms using only functional features outperformed the ones using only anatomical features (AUC=0.77 versus 0.64). The prediction accuracy slightly improved by combining both imaging modalities, and adding age and sex information provided an additional small improvement.

Additionally, we controlled for a potential confounding effect of motion (in case patients and controls moved in different amounts). We used movement parameters extracted from the functional MRI data, and extracted their mean, standard deviation, kurtosis, and skewness. We trained a logistic regression classifier on these descriptors. The prediction accuracy of this classifier on the private set was of ROC-AUC=0.61.

*M3. Effect of increasing the number of subjects*

A machine learning algorithm learns a predictive model from samples of data, here multiple subjects. Larger datasets better characterise the problem and thus lead to better prediction accuracy. We studied this improvement by varying the number of subjects (samples). Our goals were twofold: First, to estimate whether the size of the current dataset was large enough. Second, to quantify the potential gain that would be brought by increasing the number of subjects. We trained different models using numbers of subjects ranging from 500 to 1500 by steps of 250. We tested these models on a sample of 500 subjects. We generated the training and testing samples in the following manner: First, we shuffled the full dataset (i.e., public and private datasets) and randomly selected the testing sample. From the remaining samples, we took a bootstrap sample of a given size. We repeated this sampling 100 times to have estimates of the variance and the bias. Figure 2e plots the average prediction accuracy against the number of subjects (known as a “learning curve”).

The prediction accuracy steadily increased with the number of subjects. To extrapolate beyond the available number of subjects, we fit the learning curve with a suitable function: starting at 0.5 for n=0 and saturating at a fitted value with a growth in $\sqrt{n}$ (which is the convergence of the estimator). The result was $AUC = 0.5 + 0.3329\,(1 - e^{-0.0669\sqrt{n}})$. Hence, we estimated that a training dataset of 10,000 subjects will bring an AUC=0.8379. For reference, an AUC>0.8 is commonly accepted as an excellent level of discrimination (Hosmer and Lemeshow 2000).

*M4. Relative importance of different brain regions in functional MRI*

We established an ordering of importance of brain regions by assigning to each region the sum of the classifier weights on all the corresponding connections. Then, we checked the consistency of those connections across atlases and methods. For different atlases, there are different numbers of neighbours per node (a brain region). Thus, we matched the quantiles of each node summary statistic across atlases and methods to a uniform target. We then used radial basis functions to interpolate between nodes and create a continuous brain map. Figure 3a shows the spatial distribution of the average region ranking for all submissions. The information is distributed across the brain, with a slightly larger importance for regions around the precuneus. To test this spatial distribution, we removed varying proportions of the most significant brain regions (25%, 50%, 75%) and trained new models. The prediction accuracy remained high even after removing up to 50% of the brain regions (Fig. 3b).
